# Implementing an antibiogram profile to aid rational antimicrobial therapy and improving infection prevention in an urban hospital in The Gambia; strategies and lessons for low income and middle-income countries

**DOI:** 10.1101/2022.04.19.22274009

**Authors:** Saffiatou Darboe, Ruel Mirasol, Babapelumi Adejuyigbe, Abdul Khalie Muhammad, Behzad Nadjm, Annabelle de St Maurice, Tiffany L. Dogan, Buntung Ceesay, Solomon Umukoro, Uduak Okomo, Davis Nwakanma, Anna Roca, Ousman Secka, Karen Forrest, Omai B. Garner, MRCG/UCLA collaborative working group

**Affiliations:** Medical Research Council Unit The Gambia at the London School of Hygiene and Tropical Medicine; Department of Pathology and Laboratory Medicine, University of California Los Angeles, Los Angeles, California, USA; David Geffen School of Medicine, University of California, Los Angeles, Los Angeles, California, UCLA; University College London Hospital NHS Foundation Trust, London, UK; Department of Pediatrics, Division of Infectious Diseases, David Geffen School of Medicine, UCLA; Department of Clinical Epidemiology and Infection Prevention, University of California, Los Angeles, California, UCLA Health

**Author notes:** **Corresponding author:** Saffiatou Darboe, Medical Research Council Unit The Gambia at London School of Hygiene and Tropical Medicine, P.O Box 273 Banjul, The Gambia. **Alternative Corresponding Author** Dr Omai Garner.

**Keywords:** Cumulative antibiogram, antimicrobial resistance (AMR), Infection prevention and control (IPC), low- and middle-income countries (LMICs), *Escherichia coli* (*E. coli*), *Staphylococcus aureus* (*S. aureus*), *Klebsiella pnuemonaie* (*K. pneumoniae*) bacteraemia, urinary tract infection (UTI)

## Abstract

**Background:** Diagnostic microbiological capabilities remain a challenge in low- and middle-income countries resulting in major gaps. The global antimicrobial resistance burden has necessitated use of appropriate prescribing to curb the menace. This study highlights the process used to develop an antibiogram to monitor resistance at a secondary-level health facility to aid empirical clinical decision making.

**Methods:** This retrospective cross-sectional descriptive study used 3 years of cumulative data at the Medical Research Council Unit The Gambia from January 2016 to December 2018. Phenotypic data was manually imputed into WHONET and the cumulative antibiogram constructed using standardised methodologies according to CLSI M39-A4 guidelines. Pathogens were identified by standard microbiological methods and antimicrobial susceptibility testing was performed using Kirby-Bauer disc diffusion method according to CLSI M100 guidelines.

**Results:** A total of 14776 non-duplicate samples (blood cultures n=4382, urines n=4914, other miscellaneous swabs and aspirates n=2821 and n=390 respectively, sputa n=334, stools n=1463, CSF 353 and other samples n= 119) were processed of which 1163 (7.9%) were positive for clinically significant pathogens. Among the 1163 pathogens, *E. coli* (n= 315) *S. aureus* (n=232), and *K. pneumoniae* (n=96) were the leading cause of disease Overall, the susceptibility for *E. coli* and *K. pneumoniae* from all samples were: trimethoprim-sulfamethoxazole (17% and 28%), tetracycline (26% and 33%), gentamicin (72% and 46%), chloramphenicol (76 and 60%), and ciprofloxacin (69% and 59%), amoxicillin/clavulanic (77% and 54%) respectively. Extended spectrum beta-lactamase resistance was present in 23% (71/315) vs 35% (34/96) respectively. *S. aureus* susceptibility for methicillin was 99%.

**Conclusion:** This antibiogram has confirmed susceptibility to commonly used antimicrobials was higher for *E. coli* than *K. pneumoniae* with high ESBL resistance warranting surveillance. An alternative aminoglycoside with better sensitivity such as amikacin might be relevant although this was not tested and that cloxacillin remains a drug of choice for the Staphylococci.

## Introduction

Antimicrobial resistance (AMR) is recognised as a global health threat requiring appropriate containment strategies with sub-Saharan Africa (sSA) disproportionately affected [1,2]. Multiple factors transcending disciplines contribute to the development of AMR, with inappropriate use of antibiotics regarded as a major contributing factor according to the report by the WHO Global Action Plan on antimicrobial resistance [3]. Surveillance is key in understanding the epidemiology of AMR to inform appropriate public health intervention and control. Containment strategies including monitoring antibiotic usage, monitoring resistance patterns, using appropriate guidelines for treatment, and establishing effective infection prevention and control (IPC) are essential. Thus, multistakeholder collaboration to combat AMR is required. Active surveillance is paramount in monitoring resistance trends and can be an effective, data driven strategy for evidence-based decision making [4].

The cumulative antibiogram periodically summarises a healthcare facility’s antimicrobial susceptibility profiles for public health surveillance to identify changes in trends and aid clinicians in making informed empiric therapeutic choices relative to local context [5]. Furthermore, antibiograms are useful in detecting potential infectious disease outbreaks. The use of such aggregate data on local or regional resistance trends is fundamental to discern differences and changes in patterns for appropriate selection of antimicrobials for rational use and epidemiological surveillance [1,6]. Thus, diagnostics remain key in combating AMR. However, in low and middle income countries (LMIC), major gaps exist ranging from limited expertise, infrastructure, investment and lack of institutional microbiologic diagnostic capabilities [7,8]. In addition, many countries in sub-Saharan Africa have not prioritized AMR preparedness and implementation of the global action plan to tackle AMR [9,10]. Furthermore, IPC measures to reduce hospital associated infections and antimicrobial resistance surveillance are fundamental for an integrated health system strengthening approach. The understanding of the clinical implications of resistance patterns and interpretation of an antibiogram is paramount for clinical decision making. Thus, the diagnostic microbiology lab must ensure that the antibiogram is generated in collaboration with physicians, pharmacists, and other infectious diseases professionals for improved antimicrobial stewardship [11]. Additionally, antibiogram data must be generated from quality-assured clinical diagnostic microbiology laboratories using guidelines such as the Clinical Laboratory Standards Institute (CLSI) M39-A4 consensus document [12] to ensure reliability for better patient outcomes.

Considering this, the Medical Research Council Unit The Gambia at LSTHM (MRCGatLSHTM) and University of California, Los Angeles (UCLA) formed a working group to support microbiological diagnostic capabilities and implementation of an antibiogram to aid empiric therapy and IPC to improve antimicrobial stewardship. This project has provided evidence for the feasibility of harnessing collaborative technical support for implementing AMR stewardship in a sub-Saharan African country.

## Methods and Materials

### Study design and setting

This study was cross-sectional descriptive analysis of the cumulative antibiogram over a period of three years (January 2016 to December 2018), using data from the Clinical Microbiology Laboratory at the Clinical Services Department (CSD) of the MRCG at LSHTM in The Gambia. A multidisciplinary team of microbiologists, physicians, epidemiologists, and IPC specialists across the collaborating institutions met over several months to review available data.

The Gambia is the smallest mainland African country in West Africa with a population of 2.1 million with high malnutrition and decreasing malaria incidence [13–15].

The CSD has a 42-bed capacity ward and an Outpatient Department seeing approximately 50,000 adult and paediatric patients annually. The CSD provides primary and secondary-level care to sick individuals from the surrounding population with complicated cases referred to the main tertiary government hospitals. No surgical departments, obstetrics, or Intensive Care Units are present with limited neonatal admissions. It is the only facility in the Gambia with consistent routine microbiological testing capabilities for patients with suspected invasive infections.

### Microbiological procedures

Samples were processed in the diagnostic microbiology laboratory, which is both Good Clinical Laboratory Practice (GCLP; 2010) certified and ISO15189 (2015) accredited. Standard microbiological processing of samples and identification procedures were regularly performed according to standard microbiological protocols. Blood culture samples were routinely collected in BD Bactec aerobic and anaerobic adult and paediatric bottles respectively. Positive bottles were cultured on blood, chocolate and MacConkey agar plates and drops put on microscopic slides for Gram staining. Growth on plates were further characterised and identified using biochemical reagents. Pathogens considered contaminants such as coagulase negative staph, isolated >1 time and confirmed causative agents for infective endocarditis (IE) were considered. The Duke criteria for diagnosing infective endocarditis were used to determine pathogenicity of the viridans group Strep isolates. Blood and cerebrospinal fluid (CSF) samples are routinely collected for bacterial culture from patients presenting with suspected sepsis and meningitis, respectively. Microbiological data is considered community acquired as samples are collected upon presentation and long stays are rare. In addition, first isolate for patient was considered for analysis. Data is reported and stored in a local electronic medical record system (EMRS). Selective reporting of antimicrobials is yet to be introduced and specimens were anonymised for confidentiality prior to analysis.

Urine samples were considered clinically significant when growth >10^5^ of a single organism. Urines were primarily cultured on cysteine lactose electrolyte deficient (CLED) and incubated overnight at 37°C, followed by appropriate pathogen identification if growth was > 10^5^ CFUs. Other samples were cultured on appropriate agar plates such as blood, chocolate, MacConkey, sabouraud dextrose and Thayer-martin agar plates as indicated. Isolates were identified using appropriate biochemical tests; *Enterobacterales* were identified using BioMerieux API20E, other non-enteric Gram-negative were identified using API20NE. *Staphylococcus aureus* (*S. aureus)* were identified using Staphaurex Plus and mannitol salt agar testing. *Streptococcus pneumoniae* (*S. pneumoniae)* was confirmed using optochin disc, with confirmatory testing using bile solubility. *Haemophilus influenzae* (*H. influenzae)* was identified using X and V factors and serological testing. In addition, beta-haemolytic group *Streptococci* were identified using Streptex/Wellcogen serological test. Antimicrobial susceptibility patterns were determined by Kirby-Bauer disk diffusion on Mueller-Hinton agar and zone sizes interpreted according to the relevant Clinical Laboratory Standard Institute (CLSI) guidelines on antimicrobial agents [16]. Appropriate American Type Culture Collection (ATCC) controls *E. coli* 25922, *P. aeruginosa* 29835, *S. aureus* 25923 and *S. pneumoniae* 49616 were consistently used as quality control organisms for the antibiotic susceptibility testing and reagent performance verification.

### Analysis

Data were extracted from the Electronic Medical Record System (EMRS), uploaded into WHONET, cross-checked for unusual susceptibility patterns, verified as per CLSI M39 [12] and cleaned. All pathogens isolated during the period considered as individual infection episodes were included. Isolates from successive cultures from different body sites were reviewed and the first isolate included as per the CLSI guideline. Data was stratified by specimen type for blood and Urine and analysed. Heat maps were generated to visually highlight resistance patterns and bar charts to show differences in organism frequencies among samples. Tables were generated to show cumulative antibiograms for use and implement antimicrobial prescription guideline. [12].

### Ethical review and approval

The study is part of an ongoing microbiological improvement project to implement local prescribing guideline and antimicrobial stewardship at the MRCG at LSHTM.

## Results

### Samples and pathogen distribution

Over the 3-year period, a total of 14,776 different specimens were received and processed (Figure 1, flowchart), of which 1163 clinically significant organisms were isolated. Urine and blood cultures accounted for one-third (n=4914) and (n=4382) of the samples respectively.

**Figure 1:**
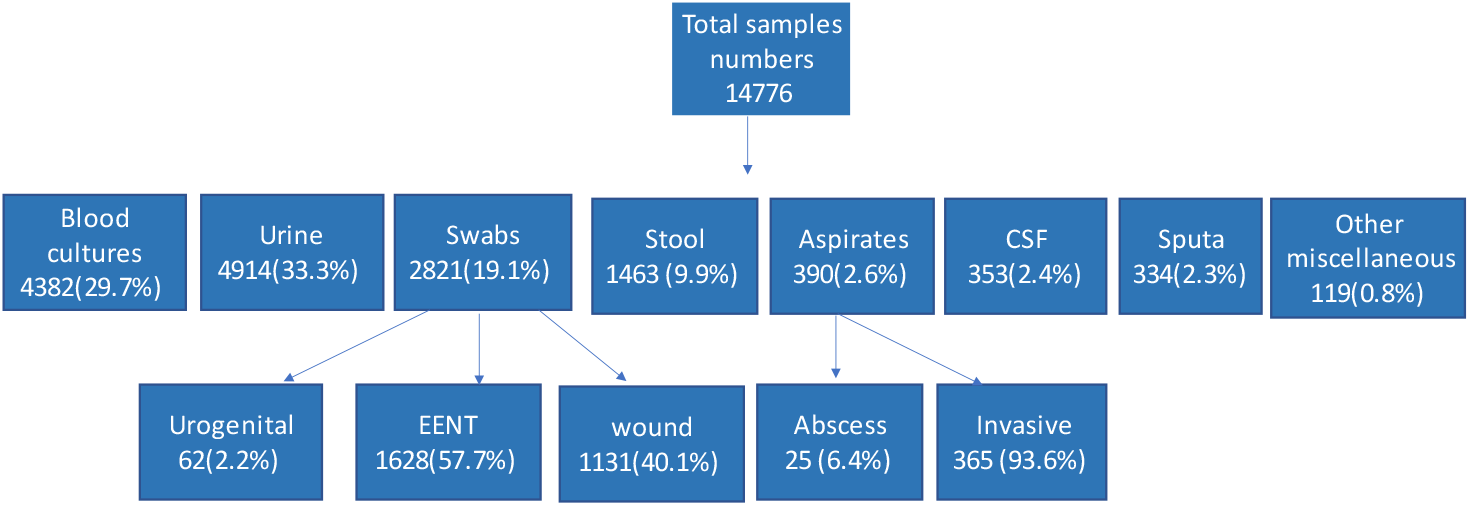
Flow chart showing distribution of the different samples

Results of 182 blood cultures, (4.2%) were considered clinically significant (Figure 2a). The leading cause of bacteraemia were *E. coli* (n=40) and *S. aureus* (n=39) responsible for 22.0% and 21.4% respectively (Figure 2b). This was followed by non-typhoidal *Salmonella* (NTS) (n=16), *S. pneumoniae* (n=15), *Enterococcus* species (n=15), and *Pseudomonas* species other than *P. aeruginosa* (n=10) (Table 1). From urine samples, 9.0% were confirmed clinically relevant (n=442) (Figure 2a). Among the causes of UTI, 55.4% were *E. coli* (n=245) and 12.7% were *K. pneumoniae* (n=56) (Figure 2b) followed by other coliform bacteria (n=31), Candida species (n=26), *Enterococcus faecalis* n=16 and *Streptococcus agalactiae* n=14 (Table 2).

**Table 1:** Susceptibility profile for the common blood culture pathogens

| Organism | NUMBER | % Susceptibility |  |  |  |  |  |  |  |  |  |  |  |  |  |  |  |
| --- | --- | --- | --- | --- | --- | --- | --- | --- | --- | --- | --- | --- | --- | --- | --- | --- | --- |
| Gram-negatives |  | PEN | AMP | SXT | CN | CHL | TET | CIP | CXM | ERY | PB | OX | FOX | CTX | CAZ | VA | AMC |
| *E. coli* | 40 | Na | 13 | 15 | 83 | 85 | 38 | 85 | 38 | Na | Na | Na | 97 | 90 | 88 | Na | 80 |
| NTS | 16* | Na | 100 | 88 | 94 | 100 | 88 | 100 | 100 | Na | Na | Na | 100 | 100 | 100 | Na | 100 |
| *Pseudomonas* species | 10* | Na | Na | 22 | 90 | 22 | 63 | 89 | 50 | Na | Na | Na | NA | Na | 100 | Na | Na |
| Gram-positives |  |  |  |  |  |  |  |  |  |  |  |  |  |  |  |  |  |
| *S. aureus* | 39 | 13 | Na | 69 | 97 | 97 | 74 | 100 | Na | 87 | Na | 100 | 100 | Na | Na | 100 | Na |
| *S. pneumoniae* | 15* | 80 | Na | 7 | Na | 93 | 67 | Na | Na | 93 | Na | 80 | Na | Na | Na | 100 | Na |
| *Enterococcus* species | 15* | 92 | 92 | Na | Na | 83 | 58 | 83 | Na | 58 | Na | Na | Na | Na | Na | 100 | Na |

\*Less than the recommended number of 30 isolates

**Table 2:**
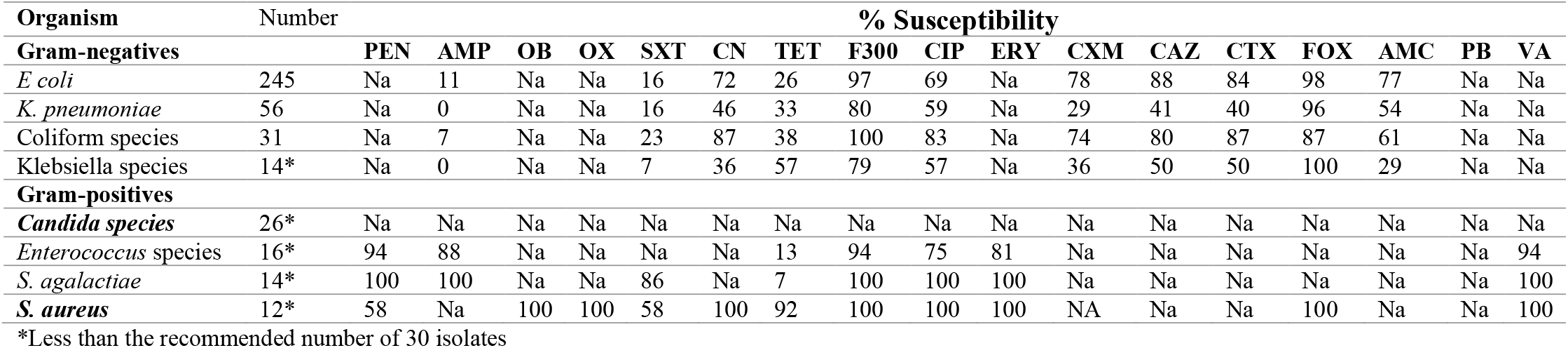
Susceptibility profile for the urine pathogens

**Figure 2a:**
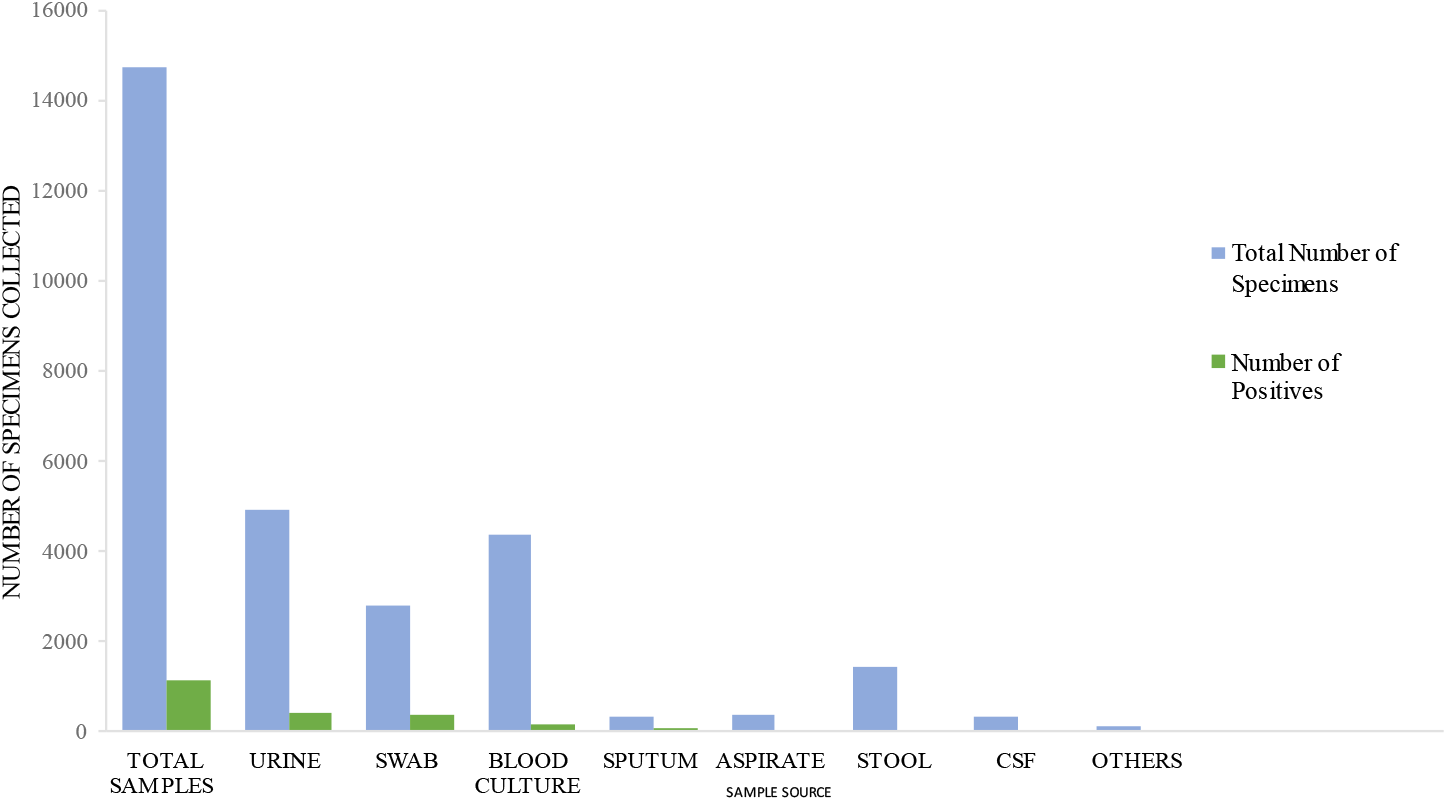
Graph showing total number of samples received against clinically significant pathogens recovered

**Figure 2b:**
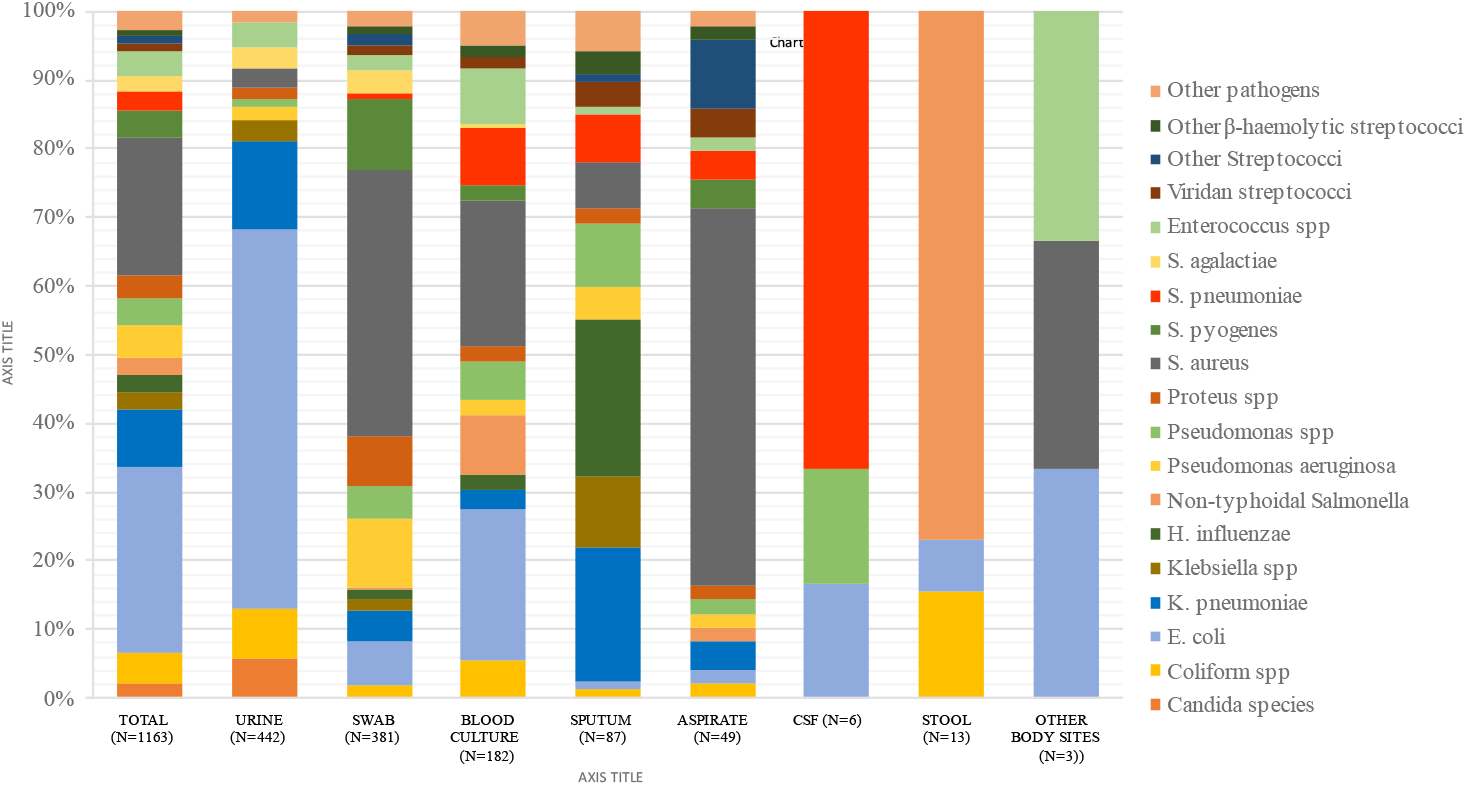
Bar chart showing proportion of pathogenic organisms by sample type

Out of 2821 swabs, a total of 381 (13.5%) were considered clinically significant. The swabs were further stratified into eye, ear, nose & throat (EENT) n=1628, skin/wound n=1131 and urogenital n=62 (Figure 1). When stratified according to swab type, 1.2% pathogens were recovered from EENT (n=19), 3.2% of pathogens were recovered from skin/wound (n=355) and 11.3% from urogenital swabs (n=7). The leading pathogens from EENT were *H. influenzae* non-type b (n=5), *K. pneumoniae* (n=4) and *S. pneumoniae* (n=4) (Supplementary figure 1). For skin/wound swab, the leading pathogens were *S. aureus* (n=148), *Pseudomonas aeruginosa* (n=39) and *S. pyogenes* (n=38). From urogenital swabs, *S. agalactiae* (4/7) and *N. gonorrhoea* (3/7) were recovered. Among the 390 aspirates, 12.6% (n=49) were considered clinically significant. They were stratified into abscess (n=25) and aspirates from invasive sterile sites (n=365). Pathogenic organisms were recovered from 96% abscesses (24/25) and 6.8% of invasive aspirates (25/365). The organisms from abscesses were 83.3% *S. aureus* (n= 20), one each of *Morganella morganii*, NTS, *Streptococcus* group F and *K. pneumoniae*. Among 25 pathogens from invasive sterile aspirates, *Streptococci* species (n=7) and S. *aureus* (n=7) were most prevalent, followed by other coliform bacteria (n=4), two *S. pneumoniae* and two *Pseudomonas* species (Supplementary figure 2). Among 334 sputa samples, 26.0% were positive for a pathogen (n=87). The leading pathogens were *H. influenzae* non-type b n=20 (23%) and *K. pneumoniae* n=17 (19.5%) (Figure 2b). Over the time period studied, only (1.7%) of 353 CSF grew pathogens (n=6) of which four were *S. pneumoniae*, one *E. coli* and one *Pseudomonas* species. Among the stool samples, 13 pathogens were isolated of which (n=10) were NTS.

Overall, *E. coli* (27.1% of the pathogens)and *S. aureus* (19.9%) and were found to be the leading cause of disease in our setting (Figure 2b) followed by *Klebsiella pneumoniae* 8.3% (n=96), *Pseudomonas aeruginosa* (n=57), other coliform bacteria (n=52), *Streptococcus pyogenes* (n=45), *Enterococcus* species (n=43), *Pseudomona*s species other than *aeruginosa* (n=43), *Proteus* species (n=41), *Streptococcus pneumoniae* (n=31) and *Klebsiella* species other than *pneumoniae* (n=30), non-typhoidal *Salmonella* (n=28) and *Streptococcus agalactiae* (n=27) (Figure 2b). *E coli* was prevalent in urine, blood and swabs whilst *S. aureus* was prevalent in swabs, blood and aspirates. In addition, *K. pneumoniae* was prevalent in urine, sputum and swabs.

### Susceptibility profiles of pathogens

Susceptibility patterns for the common pathogens found in blood and/or urinary pathogens for various antimicrobials was varied (Table 1 for blood culture and Table 2 for urinary pathogens). The major causative agents for UTI were highly susceptible to nitrofurantoin (94%). Overall, the *Enterobacterales* (except NTS) susceptibility to ampicillin was 11% for *E. coli* and 44% for *Proteus* species. Susceptibilities to 3^rd^ generation cephalosporins for *E. coli* and *K. pneumoniae* were 86% and 54%, for cephamycin 97.8% and 97.9% and for the beta-lactamase inhibitor clavulanic acid 77.2% and 65% respectively. ESBL resistance was present 23% (71/315) vs 35% (34/96) for *E. coli* and *K. pneumoniae* respectively. Susceptibilities to chloramphenicol, ciprofloxacin and gentamicin for *E. coli* were 75.7%, 72.7% and 75.9% respectively. For *K. pneumoniae* 60%, 63.5%, 53.2% and for *S. aureus* 97.3%, 98.9%, 95.2% respectively (Figure 3). Remarkably, susceptibility for methicillin for *S. aureus* was very high 99.4% but low for penicillin 20.2%. *S. pneumoniae* and *H. influenzae* were susceptible to penicillin 77.4% and ampicillin 100% respectively but low for trimethoprim-sulfamethoxazole (0%). Penicillin susceptibility for *Enterococcus* was 81.4%. *S. agalactiae* and *S. pyogenes* susceptibilities were 100% susceptible for penicillin but for tetracycline 33.3% and 29.6.% respectively. These susceptibility profiles are shown in the heatmap depicting increasing resistance with from purple to red (Figure 3)

**Figure 3:**
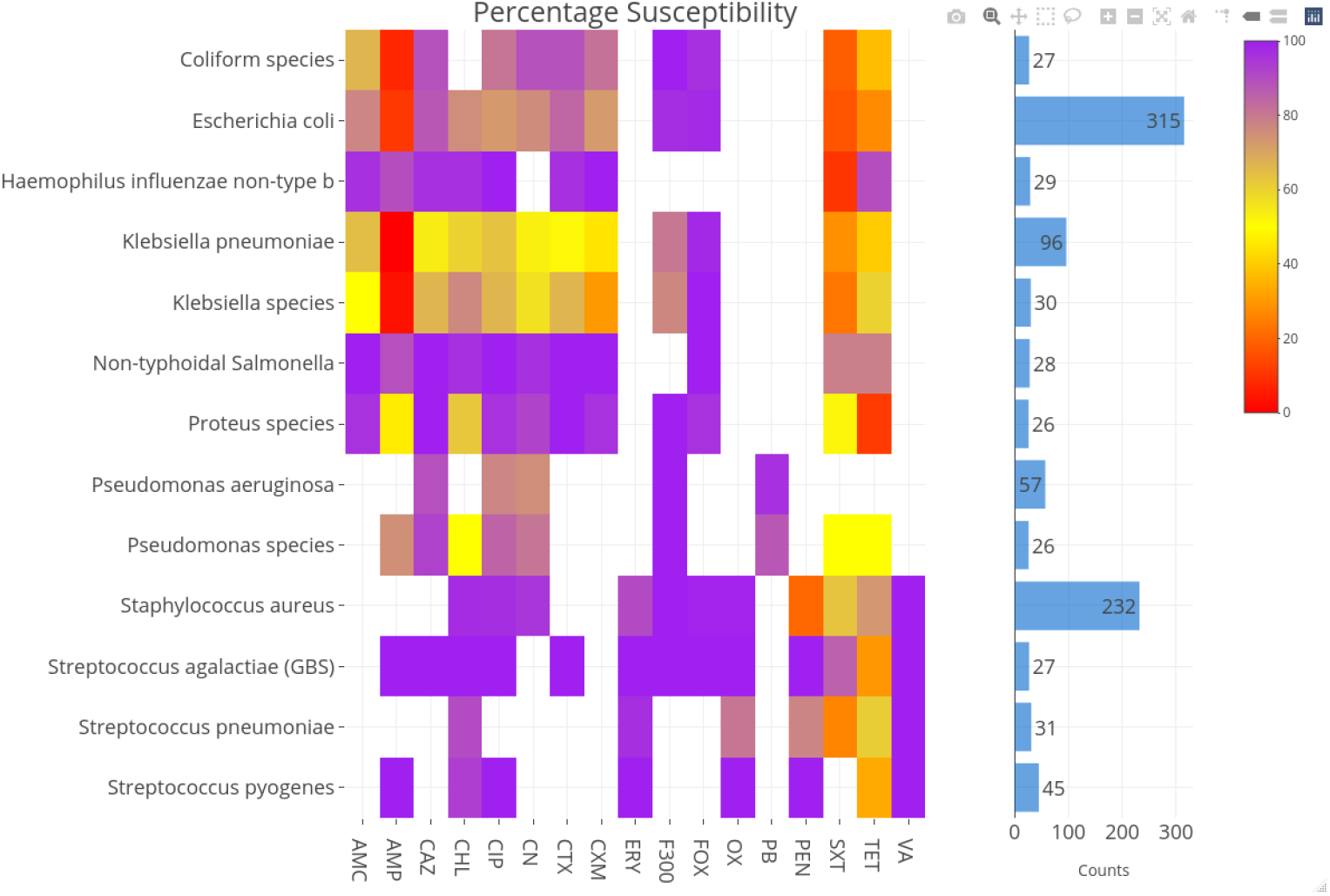
Heatmap showing the intensity of resistance patterns for the different pathogens. Purple depicting susceptibility and increasing resistance shown from yellow to red

## Discussion

This study provides an antibiogram profile from the diagnostic microbiology laboratory for clinical samples over a three-year period in The Gambia to aid in empirical clinical decision making. This is part of a collaborative network between the MRCG, and UCLA set out to improve microbiological laboratory capacity and improve IPC. Diagnostic microbiological capabilities with standardised methodologies and expertise for detection and management of AMR are often scarce in many LMICs [6,17]. The global action plan on antimicrobial resistance highlights inappropriate prescribing as one of the main drivers of AMR [3], thus, the implementation of this antibiogram is paramount in understanding resistance dynamics to aid rational local prescribing. The antibiogram is particularly useful when the infecting organism is known prior to susceptibility patterns. With the global spread of antimicrobial resistance[2,18], it is a useful tool to monitor local changes to provide evidence-base data for local prescribing guidelines. However, it is important that ways of disseminating the information to all prescribers are facilitated for easy access in prescribing areas. It is also important that users are educated in its effective use and effect on patient outcome [19] and thus this study facilitated that in a local hospital.

In this study, *E. coli* and *S. aureus* were found to be the most prevalent cause of all disease and in particular bacteraemia. It is important to note that *S. aureus* remains a prevalent cause of bacteraemia [15,20] as seen in other parts of globe [21,22]. However, *E. coli* is for the first time being reported in the top two bacteraemia pathogens, taking over from *S. pneumoniae* as reported previously [15,20]. In addition, it remains the prevalent cause of UTIs. The reduced importance of *S. pneumoniae* as a cause of bacteraemia in The Gambia is attributed to the increasing impact of pneumococcal conjugate vaccines over the years after a higher prevalence of children have been vaccinated [23,24]. The vaccine effectiveness studies done in The Gambia have shown both substantial reduction in incidence of pneumococcal bacteraemia among young children [24] and in pneumonia hospitalisation [25]. Invasive NTS remains a major cause of bacteraemia albeit with low resistance found in this setting. Notwithstanding, the geographical differences noted for NTS serovars and resistance patterns in previous studies ([26,27] warrant advances in surveillance and monitoring to provide information on prescribing.

The coagulase negative *Staphylococci* and viridan group *Streptococci* were mainly considered contaminant unless in patients with underlying conditions such as infective endocarditis, malignancy and neutropenia after meeting criteria for clinical relevance [28]. It is particularly important to state that quality improvement to reduce contaminant rates and improved collection techniques to increase yield in blood cultures is also ongoing.

The urine samples harboured the most common bacterial isolates with *E. coli, K. pneumoniae* and other coliforms as the leading cause of community acquired UTI. Patients were not stratified by age, sex, symptoms or with co-morbidities as this was beyond the scope of this study. A more robust study considering these factors is warranted considering it’s a significant cause of morbidity and burden in The Gambia [29,30]. It is important to note that the use of indwelling catheters is uncommon in this setting. However, the overall causes of UTIs in The Gambia are of Gram-negative origin as in other studies [31–33]. *Enterococcus faecalis* and *Streptococcus agalactiae* was also found to cause disease.

The susceptibilities for *E. coli* were similar for both disease syndromes except for ciprofloxacin which was comparatively lower for urinary tract strains (69% vs 85%) than for blood strains. This warrants further investigation into the genomic epidemiology of this important invasive pathogen. Although susceptibility profiles for *E. coli* and other *Enterobacterales* remain high except for *K. pneumoniae*, lower susceptibility is reported in this study than previously for ciprofloxacin, gentamicin, chloramphenicol, and 3^rd^ generation cephalosporins [15]. However, the lower ciprofloxacin susceptibility profile seen for *E. coli* in UTI compared to bacteraemia deserves further investigation and characterisation. Multidrug resistance was evident especially for ampicillin, tetracycline and trimethoprim-sulfamethoxazole. Both Gram-negative and Gram-positive pathogens had low susceptibilities profiles for trimethoprim-sulfamethoxazole and tetracycline making these ineffective in our setting. It is worth highlighting that carbapenem resistance is yet to be established routinely. Importantly, in the era of the decline of pneumococcal meningitis and bacteraemia ([15,24,25], we continue to find increasing resistance to penicillin in this study highlighting the need for surveillance.

The WHO recommended empiric drug of choice for sepsis remain for ampicillin and gentamicin. This data therefore provides evidence for the modification of this combination to include cloxacillin and amakicin. *S. aureus* remains susceptible to cefoxitin, the proxy for methicillin and show that cloxacillin remains effective in our setting as previously described [15,34–36]. Penicillin, tetracycline, and trimethoprim-sulfamethoxazole susceptibility however was low for most pathogens including *S. aureus*. These drugs are widely available to the public and misused due to lack of regulation. The emerging penicillin resistance warrant further surveillance and highlights the need for improved microbiologic diagnostic capabilities and local antibiogram for appropriate antimicrobials. However, implementation of appropriate IPC interventions and adherence to guidelines with supporting diagnostic evidence remains a major challenge in sub-Saharan Africa [37].

In the advent of AMR, data on local context could be used to inform therapeutic guidelines and improve prescribing and impact on stewardship. To overcome the challenges of AMR, it is important to employ a combination of strategies including improving sanitation to reduce infections episodes, use of guidelines for standardized diagnostic methodologies, and improved expertise and knowledge in understanding the threat posed by this silent pandemic. In additions, reducing infections through improving hygiene and strengthening IPC in hospitals is a key intervention strategy in curbing the spread of resistance. In resource limited settings where there has been an increase in empiric treatment with WHO ‘reserve’antibiotics [3], the knowledge and use of a local antibiogram and investing in infection prevention control are paramount for the management of AMR. In addition, the key involvement of the microbiology leads with insight into the clinical relevance of the results generated has added value into this interpretation. It is important that laboratory staff are aware of the public health and clinical significance of their findings [38]. The inclusion and involvement of physicians in the process was useful in understanding of the interpretation of the antibiogram and facilitated implementation in routine prescribing as recommended [39].

This study highlights the feasibility of implementing AMR containment strategies to improve prescribing guideline in a LMIC and has provided evidence that multistakeholder collaborative effort could improve antimicrobial stewardships across borders. The implementation of the cumulative antibiogram using standardised methodology for the first time is a great opportunity to further disseminate the knowledge and skills across the entire country. This has important implications for antimicrobial stewardship policy. There are several limitations in this study that may hinder its general applicability. First, although the first isolates for patient was included in the analysis, it did not reveal if sample was collected at point of admission or during admission. Hence, we could not reliably identify a potential hospital associated infection. Second, the data was not stratified by age and risk factors. Thirdly, time of sample collection was not documented. Notwithstanding, this antibiogram data has shown high susceptibility profiles for pathogens of public health significance. We have also demonstrated that engagement and multistakeholder collaboration can be harnessed to solve global challenges such as AMR. We have shown that common antimicrobials are still effective in this setting but importantly, evidence for an improvement in combination therapy is warranted locally.

## Data Availability

All data produced in the present study are available upon reasonable request to the authors

## Acknowledgement

We thank staff of Clinical Services Department and Clinical Laboratories for their support, comments, and suggestions.

## Contributors

SD, DN, KF and OG designed the study. SD and OG wrote the draft manuscript. SD did the laboratory analysis and cleaned the data; SD, BA, AKM, RM and OG analysed the data prepared the tables and figures; BN and KF provided clinical care to the patients and facilitated implementation of the antibiogram into clinical practice; All authors critically reviewed and contributed to the manuscript.

**\*** Other members of the MRCG/UCLA collaborative working group

Usman Nurudeen Ikumapayi

Rasheed Salaudeen

Mamina Bojang

Patrick Okot

Edwin Kamau

## Figure legends and tables

**Supplementary figure 1:**
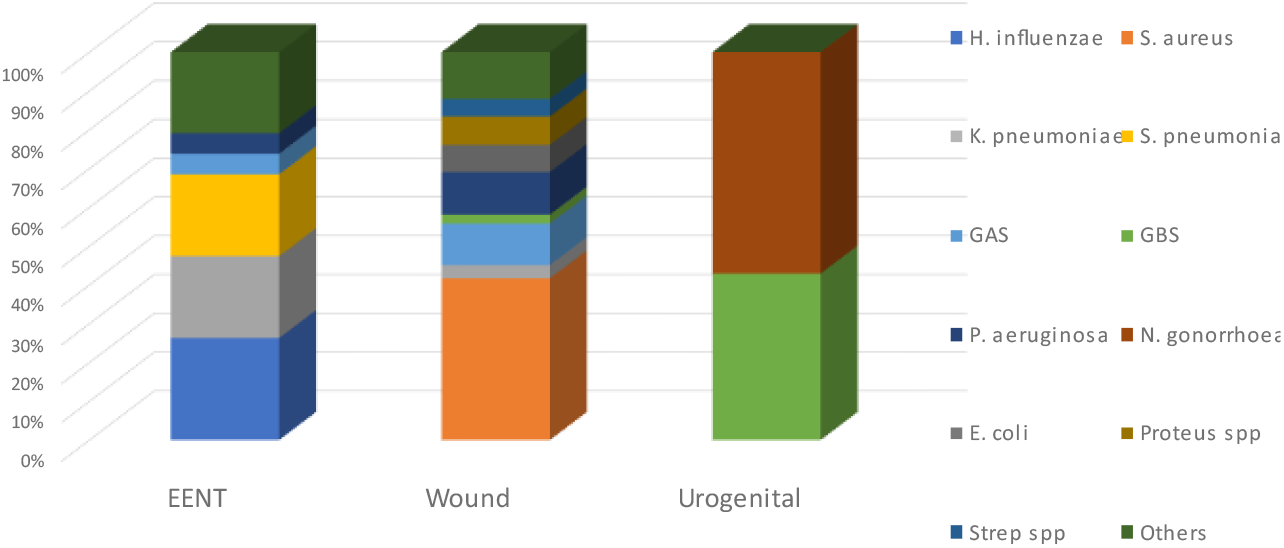
Bar chart showing the percentage of pathogens recovered from the various swabs

**Supplementary figure 2:**
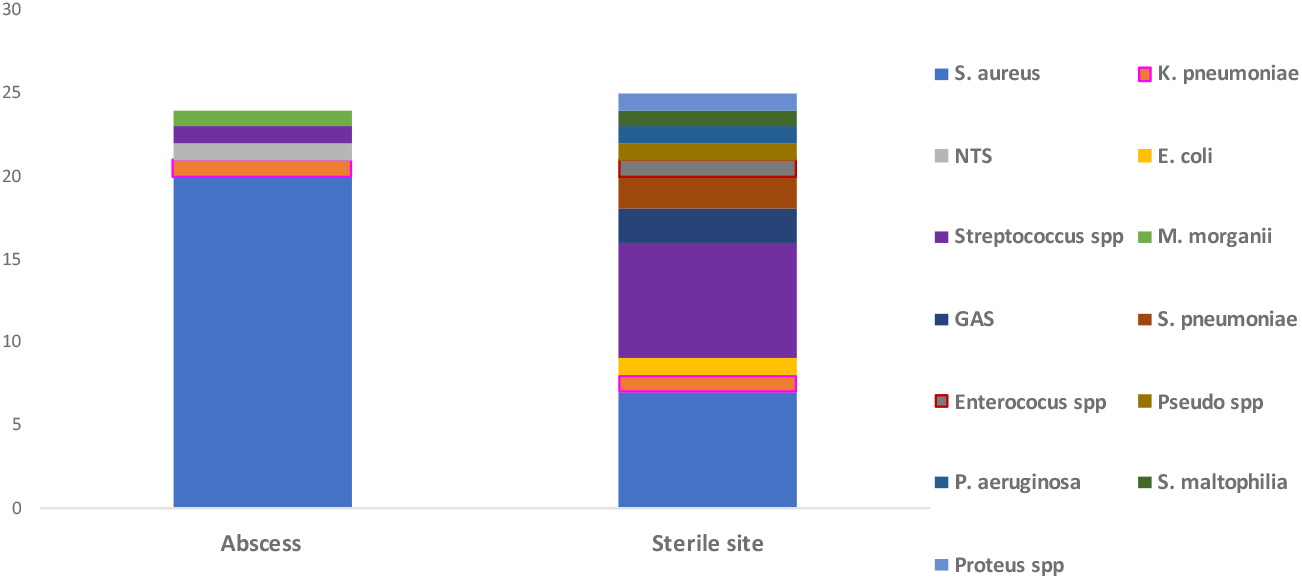
Bar chart showing the absolute numbers for the pathogens recovered from the various aspirates

